# A Glioma Stem Cell-Associated Signature Predicts Survival Across Adult and Pediatric High-Grade Gliomas and Reveals the FAM86B1/FAM86B2 Axis

**DOI:** 10.1101/2025.10.15.25338073

**Authors:** Qiqi Xie, Bokai Wang, Qin Ma, Jia Shen

## Abstract

High-grade gliomas (HGGs), including adult glioblastoma (GBM) and pediatric diffuse intrinsic pontine gliomas (DIPGs), are sustained by glioma stem cells (GSCs) that drive tumor initiation, therapeutic resistance, and recurrence. Although numerous prognostic models have been proposed, few are directly grounded in the core biology of GSCs across both adult and pediatric HGGs. In this study, we defined a GSC-associated gene signature by integrating transcriptomic profiles from patient-derived GSCs and their differentiated counterparts using in-house DIPG13 RNA-seq and the public GSE54791 dataset. The biological relevance of this signature was supported by functional enrichment and protein-protein interaction analyses. To assess its prognostic value, we applied machine learning-based modeling in a large training cohort (Chinese Glioma Genome Atlas, CGGA) and validated the resulting model across three independent datasets (Gravendeel, Rembrandt, and an integrated pediatric HGG cohort), demonstrating consistent predictive performance. To enhance clinical applicability, we developed a nomogram integrating the gene signature-derived risk score with key clinical factors, including age, race, and radiation therapy status, enabling individualized survival prediction. To further support the biological basis of the model, we experimentally examined *FAM86B1*, one of the five genes in the final signature and a gene not previously characterized in glioma biology, and found that the closely related FAM86B1/FAM86B2 axis was enriched in GSCs and overexpressed in GBM tissues, while suppression of this axis impaired GSC maintenance in both GBM and DIPG GSCs. Collectively, this study establishes a biologically grounded, GSC-centered prognostic model for HGG that improves patient stratification and may inform personalized therapeutic strategies.

## 1. Introduction

High-grade gliomas (HGGs) represent a devastating group of primary brain tumors that arise in both adult and pediatric populations (1, 2). Classified by the World Health Organization (WHO) as grade 3 or 4 neoplasms, they include glioblastoma (GBM) in adults and pediatric HGGs (pHGGs), with diffuse intrinsic pontine gliomas (DIPGs) representing the most lethal subtype. For patients with GBM, median survival rarely exceeds 15 months (3), while children with DIPG, an inoperable brainstem tumor typically diagnosed between the ages of 5 and 10, face a median survival of less than one year (4). The uniformly poor outcomes associated with these malignancies underscore the urgent need for improved prognostic tools and therapeutic strategies.

Glioma stem cells (GSCs) are a major driver of therapeutic resistance and recurrence in both adult and pediatric HGGs (5, 6). These cells possess robust self-renewal capacity, initiate and sustain tumor growth, and exhibit resistance to conventional therapies. Controlled by unique transcriptional programs, GSCs are molecularly and functionally distinct from the bulk tumor population (7–9). Despite their recognized biological importance, however, the prognostic relevance of GSC-associated molecular features remains insufficiently defined. Most existing prognostic models are constructed from bulk tumor transcriptomes, where signals from the small yet clinically consequential GSC population are frequently masked (10–12). Furthermore, models developed for specific subtypes such as GBM often fail to achieve robustness and generalizability when applied across diverse HGG cohorts, particularly in extending from adult to pediatric cases, limiting their broader clinical applicability.

To address these limitations, we developed a prognostic model grounded in a transcriptional signature that defines the GSC state across adult and pediatric HGGs. Built from in-house and public datasets and validated in four independent cohorts, the machine learning-derived model consistently outperformed existing signatures. Integrated with clinical variables into a nomogram, it provides a robust tool for individualized survival prediction and underscores the central role of cancer stemness in HGG outcomes, further supported by experimental validation of the FAM86B1/FAM86B2 axis in GSC maintenance.

## 2. Results

### 2.1. Identification of a core set of GSC–associated genes in HGGs

To define a robust set of GSC-associated genes in HGGs, we integrated GSC transcriptomic data from both in-house and public datasets (Fig. 1A). RNA-seq was performed on the pediatric HGG cell line DIPG13 cultured under stem cell–maintaining versus differentiation conditions (Fig. 1B), and these data were analyzed together with an independent GBM dataset (GSE54791) containing matched GSC and differentiated DGC pairs. Differential expression analysis identified genes consistently upregulated in GSCs across both datasets, and their intersection yielded a core set of 603 GSC-associated genes (Supplementary Table 1; Fig. 2A). Functional relevance of these genes was assessed through PPI network analysis, which highlighted known stemness regulators such as SOX2 and OLIG1/2 as top hub genes (Fig. 2B and C), supporting the validity of the approach. GO (Fig. 2D) and KEGG pathway analyses (Fig. 2E) further demonstrated that these genes are enriched in key biological processes related to GSC biology, including neurogenesis, RNA splicing, and cancer signaling.

**Fig. 1.**
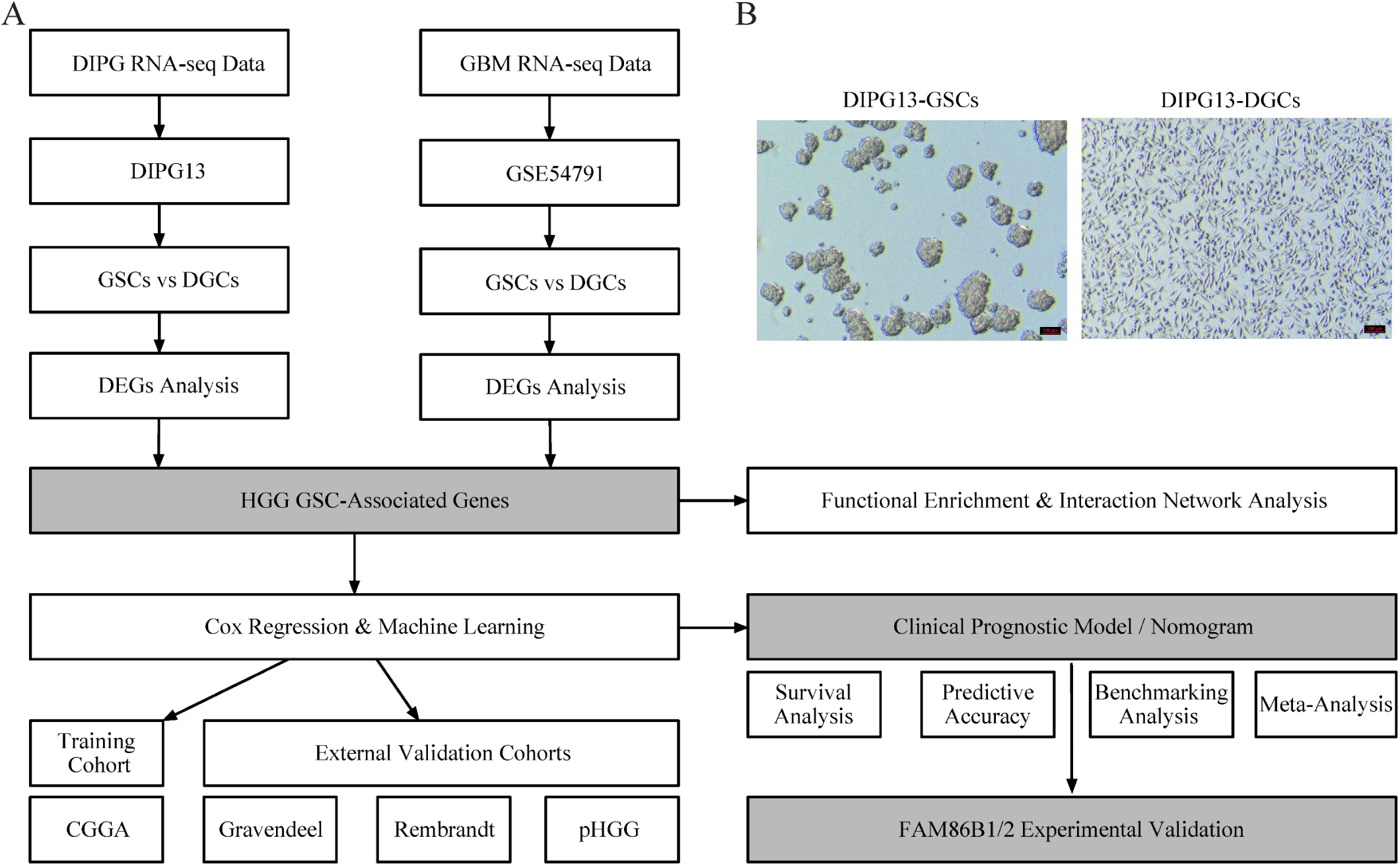
Schematic overview of the study design. (A) RNA-seq datasets from DIPG13 (in-house, pediatric HGG) and GSE54791 (adult GBM, HGG) were analyzed to identify differentially expressed genes (DEGs) between glioma stem cells (GSCs) and differentiated glioma cells (DGCs). The intersecting DEGs defined a core set of HGG GSC-associated genes, which were subjected to functional enrichment, network analysis, Cox regression, and 135 machine learning algorithms to construct prognostic models. Model performance was trained in the CGGA cohort and validated in three independent cohorts (Gravendeel, Rembrandt, and pHGG). Subsequent survival, predictive accuracy, benchmarking, and meta-analyses established a clinically applicable prognostic model and nomogram. The final model also guided experimental validation of the FAM86B1/2 axis in GSC maintenance. (B) Representative images of DIPG13 cells under stem cell conditions (left) and following serum-induced differentiation (10% FBS, right). DIPG13 cells displayed neurosphere morphology in stem cell medium and adopted an adherent morphology upon differentiation. Scale bars, 100 μm.

**Fig. 2.**
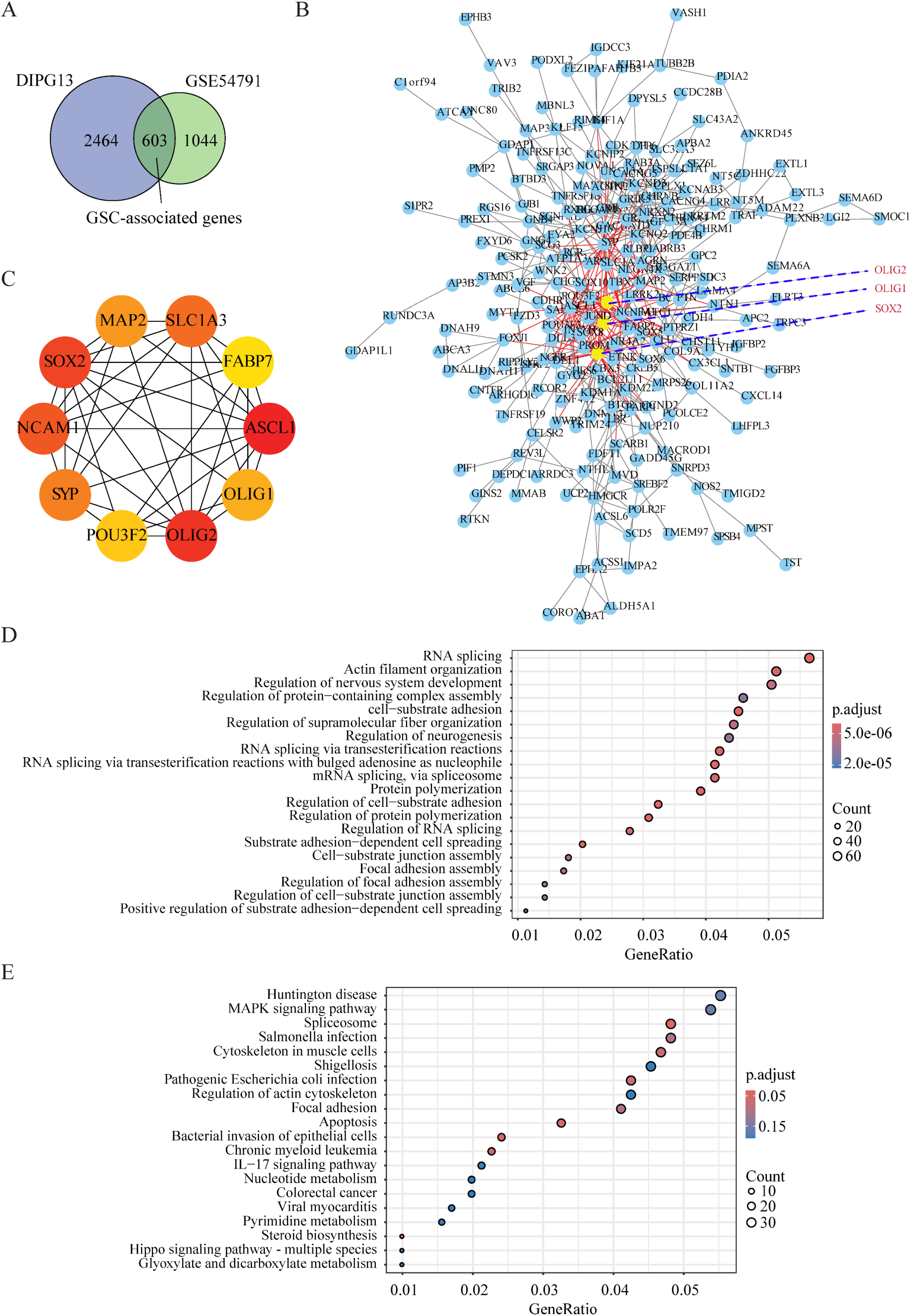
Functional characterization of HGG GSC-associated genes. (A) Venn diagram showing overlap of DEGs between GSCs and DGCs in DIPG13 and GSE54791 datasets. The 603 shared DEGs were defined as the core HGG GSC-associated genes. (B) PPI network of these 603 genes generated using STRING, with key regulators such as SOX2 and OLIG1/2 highlighted. (C) Top functional module from the PPI network identified by MCODE, highlighting 10 hub genes. (D) GO enrichment analysis for biological processes. (E) KEGG pathway enrichment analysis. In (D) and (E), dot size reflects gene counts and color indicates adjusted p values.

### 2.2. Prognostic value of the GSC-associated gene signature

To evaluate the prognostic value of the GSC-associated gene signature, we implemented a large-scale modeling strategy that generated 135 machine learning pipelines within the CGGA training cohort (Fig. 3A). Each pipeline combined one of ten feature selection methods with several survival learning algorithms, and model performance was assessed using the C-index across the CGGA training set and three independent validation cohorts (Gravendeel, Rembrandt, and the integrated pediatric HGG cohort). Among all models, the combination of forward stepwise Cox regression (StepCox[forward]) and RSF achieved the highest and most consistent C-index across datasets. Using this model, patients were stratified into high-and low-risk groups based on the median risk score derived from the training cohort. Kaplan–Meier survival analyses showed that high-risk patients had significantly shorter overall survival than those in the low-risk group, with hazard ratios ranging from 2.36 in the Gravendeel cohort (95% CI: 1.77–3.13, p < 0.001) to 4.27 in the CGGA cohort (95% CI: 3.40–5.58, p < 0.001) (Fig. 3B). Time-dependent ROC analyses further confirmed the predictive capacity of the model, with AUC values exceeding 0.80 in the CGGA cohort and above 0.70 in the Gravendeel, Rembrandt, and pediatric HGG cohorts (Fig. 3C and 4).

**Fig. 3.**
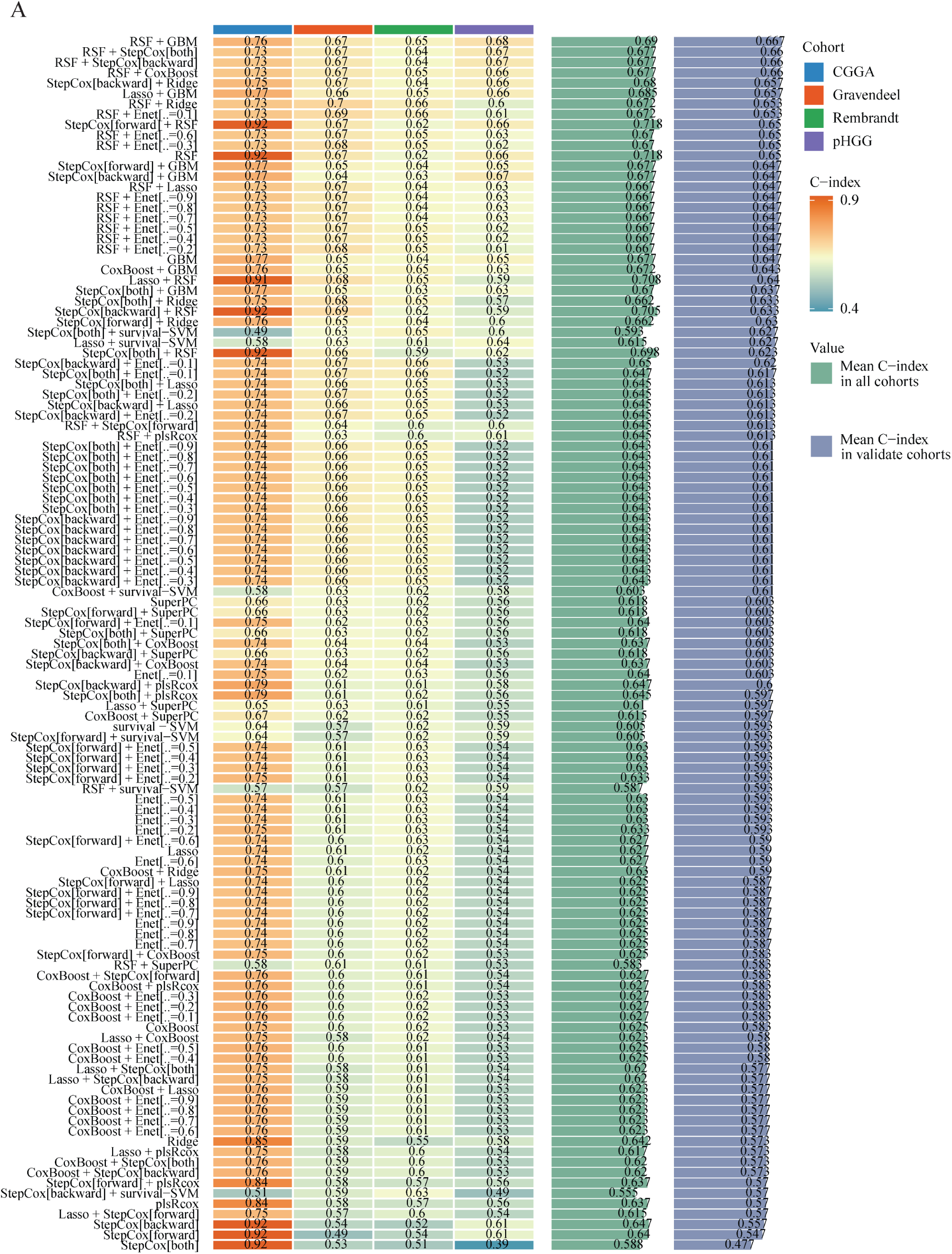
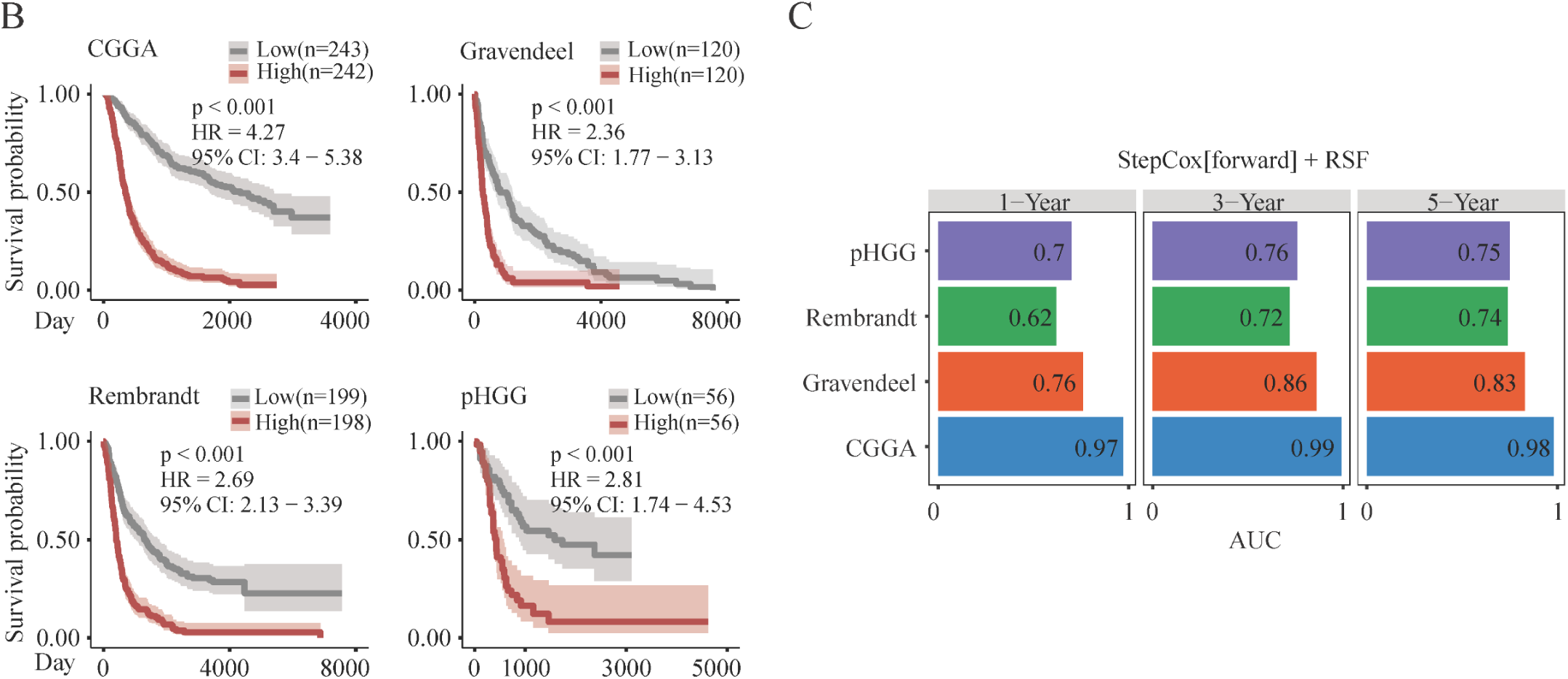
Development of the optimal machine learning–based prognostic model. (A) Heatmap summarizing the performance of 135 machine learning algorithm combinations across four HGG cohorts (CGGA, Gravendeel, Rembrandt, and pHGG), measured by C-index. Bars on the right indicate mean C-index across all cohorts and within validation cohorts. (B) Kaplan–Meier survival curves for the RSF model, showing significantly worse outcomes in high-risk versus low-risk patients (log-rank p < 0.001). Hazard ratios (HR) with 95% confidence intervals (CI) are indicated. (C) Time-dependent AUCs at 1-, 3-, and 5-years for the StepCox[forward] + RSF model.

**Fig. 4.**
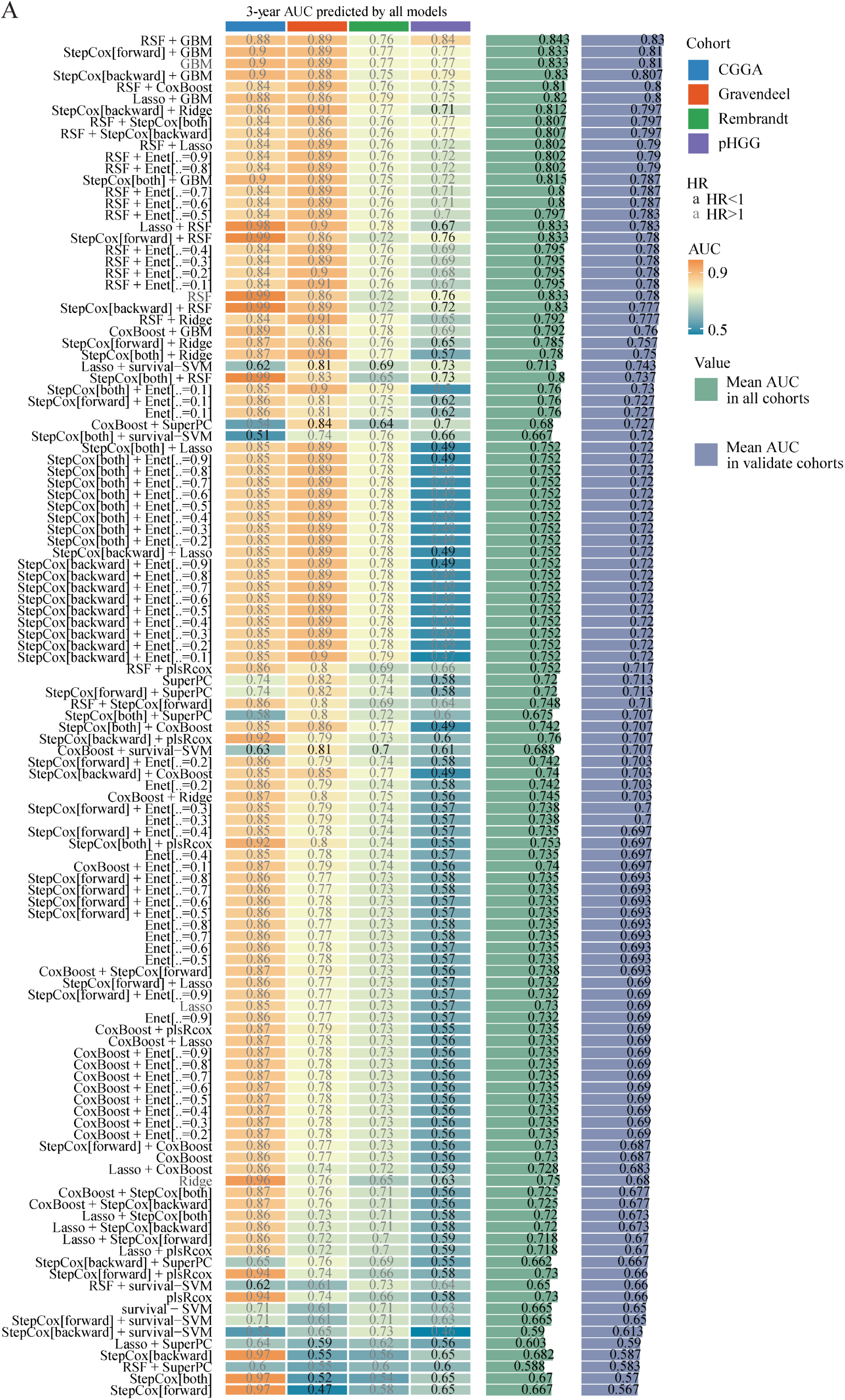
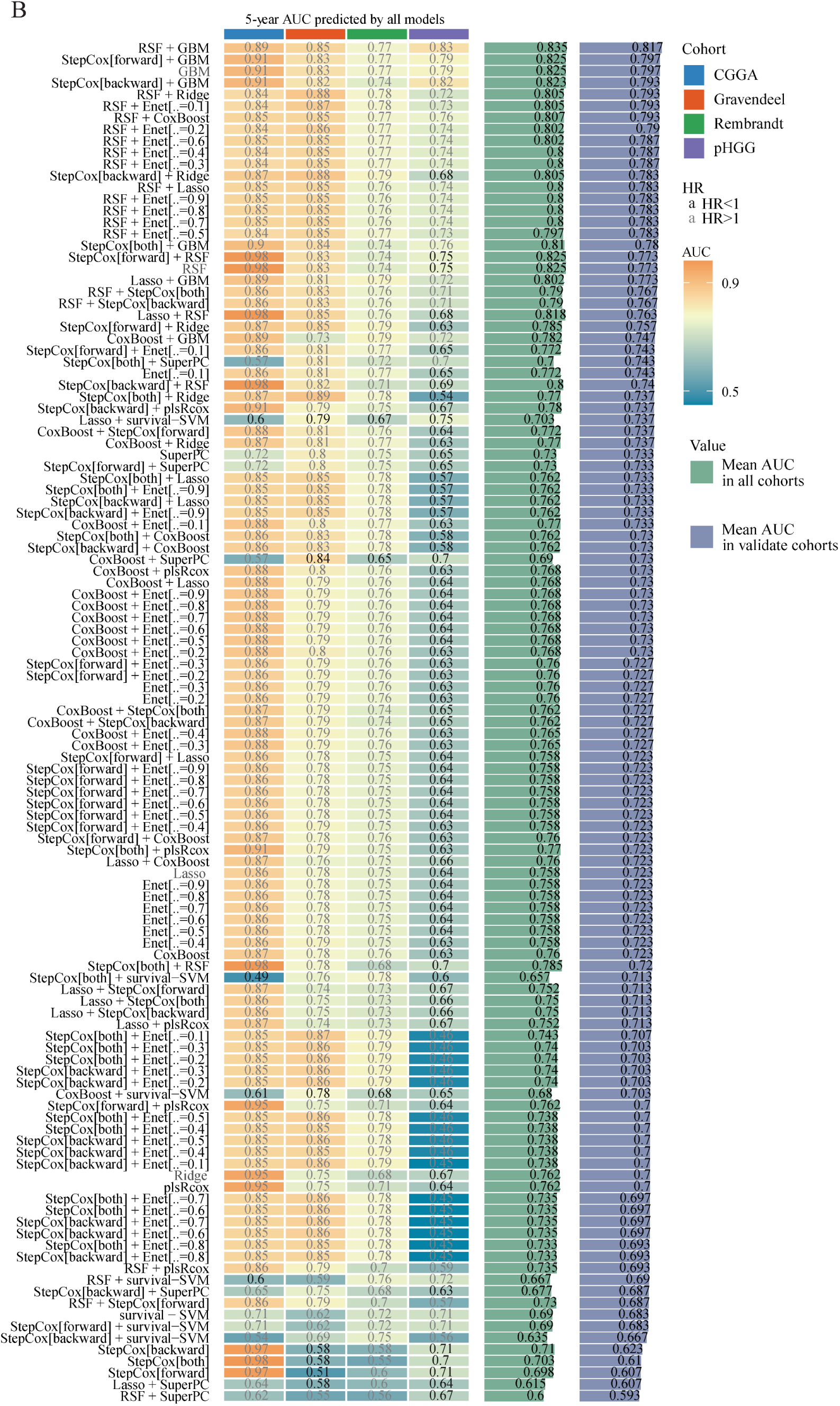
Comparative evaluation of predictive accuracy across candidate models. (A–B) Heatmaps comparing the predictive performance of all 135 candidate models across four HGG cohorts (CGGA, Gravendeel, Rembrandt, and pHGG). Performance was evaluated using time-dependent AUC at (A) 3 years and (B) 5 years. Each heatmap column represents an individual model, displaying its corresponding HR, cohort-specific AUC values, and mean AUC across cohorts. Right-side bars summarize the mean AUC values within all cohorts and specifically within the validation cohorts.

To further validate its robustness, the GSC-based prognostic model was compared with multiple previously published glioma signatures across all four datasets. Using both AUC and C-index as evaluation metrics, our model consistently ranked among the top performers, achieving the highest predictive accuracy in the CGGA cohort and comparable or superior performance in the Gravendeel and pediatric HGG cohorts (Fig. 5 and 6). Time-dependent ROC analyses across all datasets confirmed high discrimination for 1-, 3-, and 5-year overall survival predictions (Fig. 7A–C). Univariate Cox regression analysis also demonstrated that the GSC-derived risk score (GSRScore) was a significant independent predictor of mortality in CGGA (HR = 40.10, 95% CI: 26.67–60.27, p < 0.001), Gravendeel (HR = 2.65, 95% CI: 2.00–3.51, p < 0.001), Rembrandt (HR = 2.14, 95% CI: 1.70–2.68, p < 0.001), and the pediatric HGG cohort (HR = 1.40, 95% CI: 1.36–1.76, p < 0.001) (Fig. 7D). A pooled meta-analysis yielded a summary HR of 3.08 (95% CI: 0.48–19.89) under the random-effects model and 2.91 (95% CI: 2.50–3.40) under the fixed-effects model (Fig. 7D). Collectively, these results demonstrate that the GSC-associated gene signature provides robust and consistent prognostic value across both adult and pediatric HGG datasets.

**Fig. 5.**
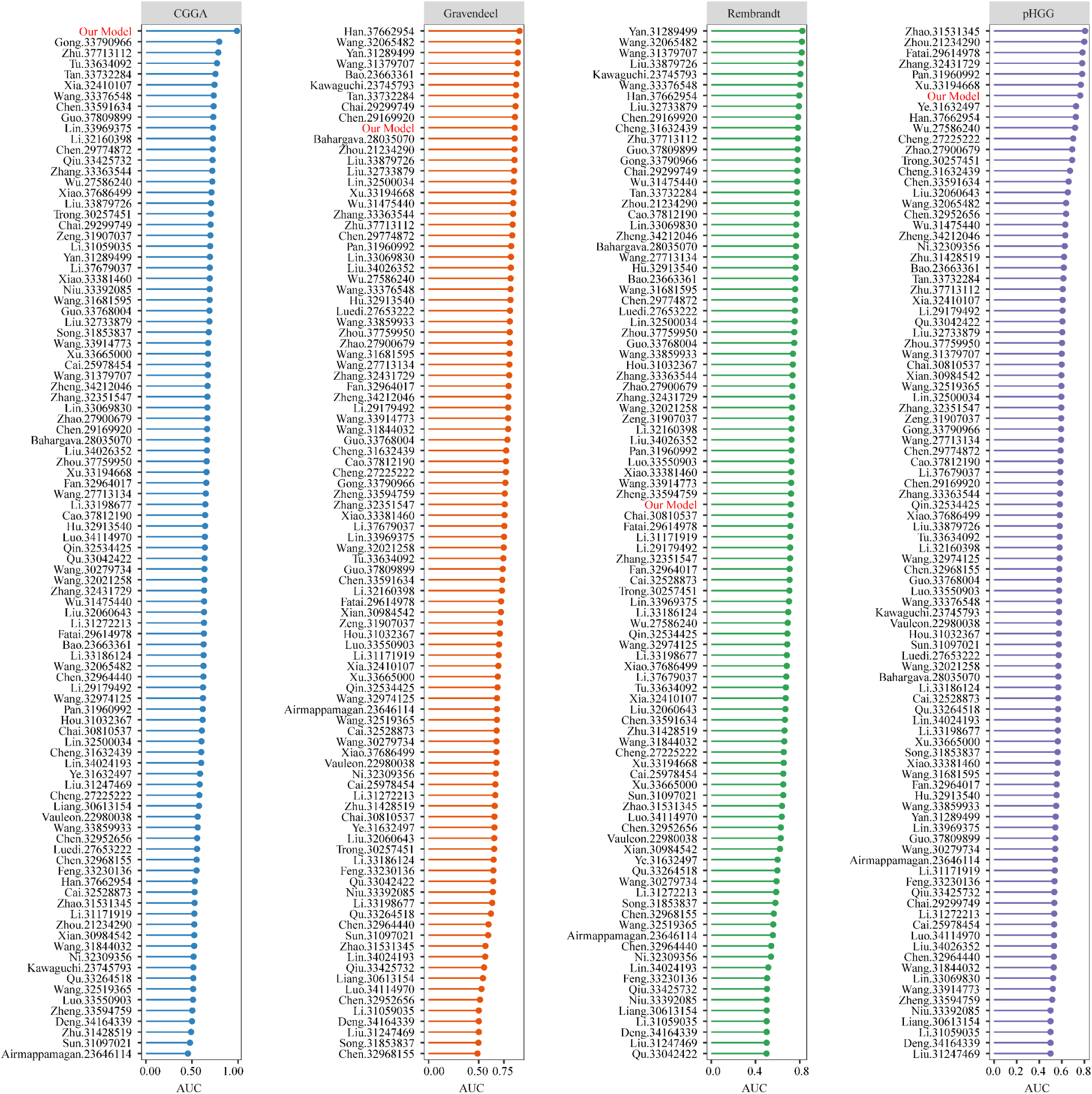
Benchmarking of the proposed prognostic model against published signatures using AUC. Dot plots comparing the AUC of the newly developed prognostic model (highlighted in red) with a comprehensive set of previously published prognostic gene signatures. Analyses were performed independently across four HGG cohorts: CGGA, Gravendeel, Rembrandt, and pHGG. Each dot represents the AUC performance of a signature within a cohort.

**Fig. 6.**
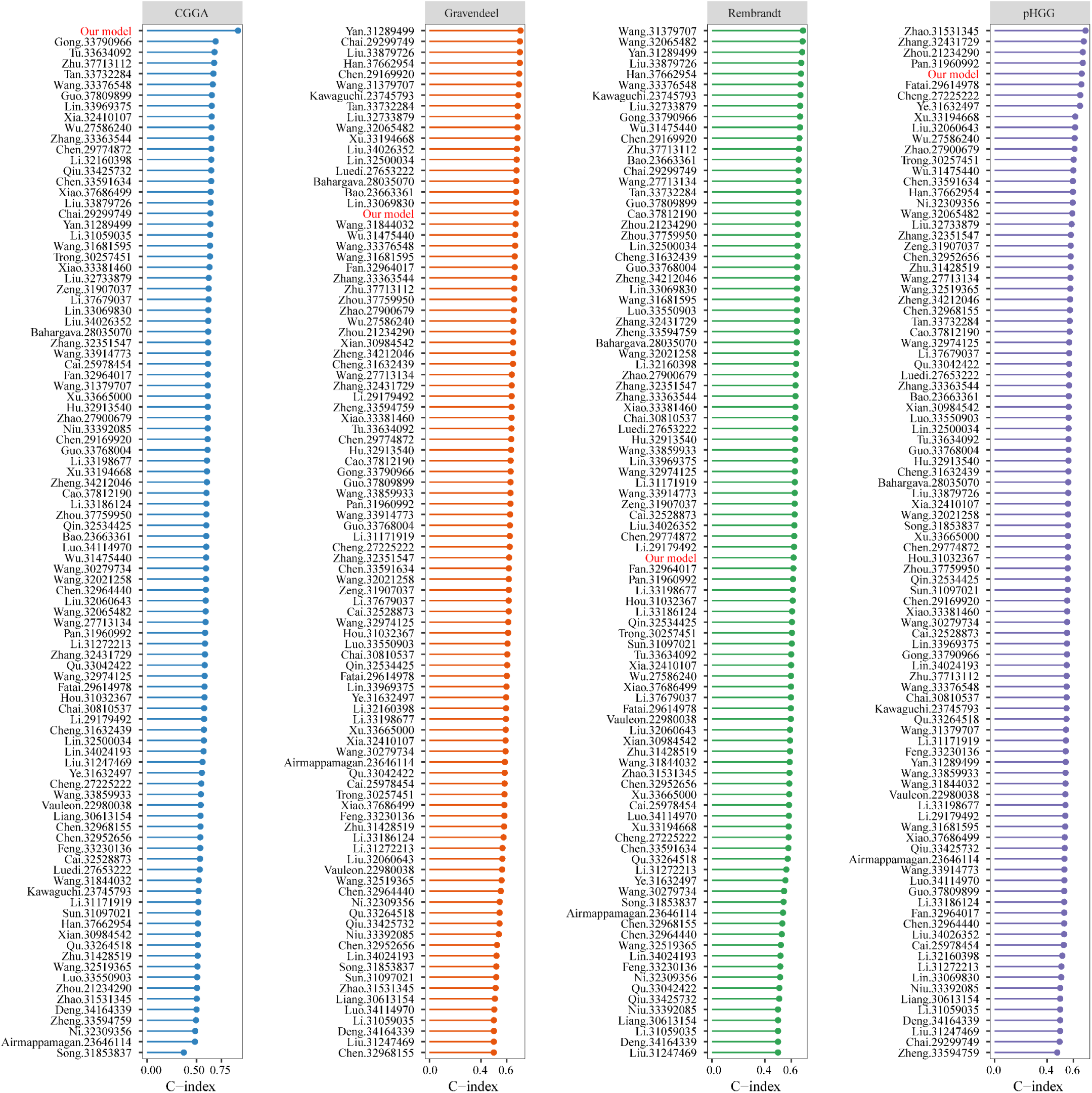
Benchmarking of the proposed prognostic model against published signatures using C-index. Dot plots illustrating the C-index of the new prognostic model (highlighted in red) compared with a wide range of previously published prognostic gene signatures. Analyses were conducted across the CGGA, Gravendeel, Rembrandt, and pHGG cohorts. Each dot indicates the C-index for an individual signature within a given cohort.

**Fig. 7.**
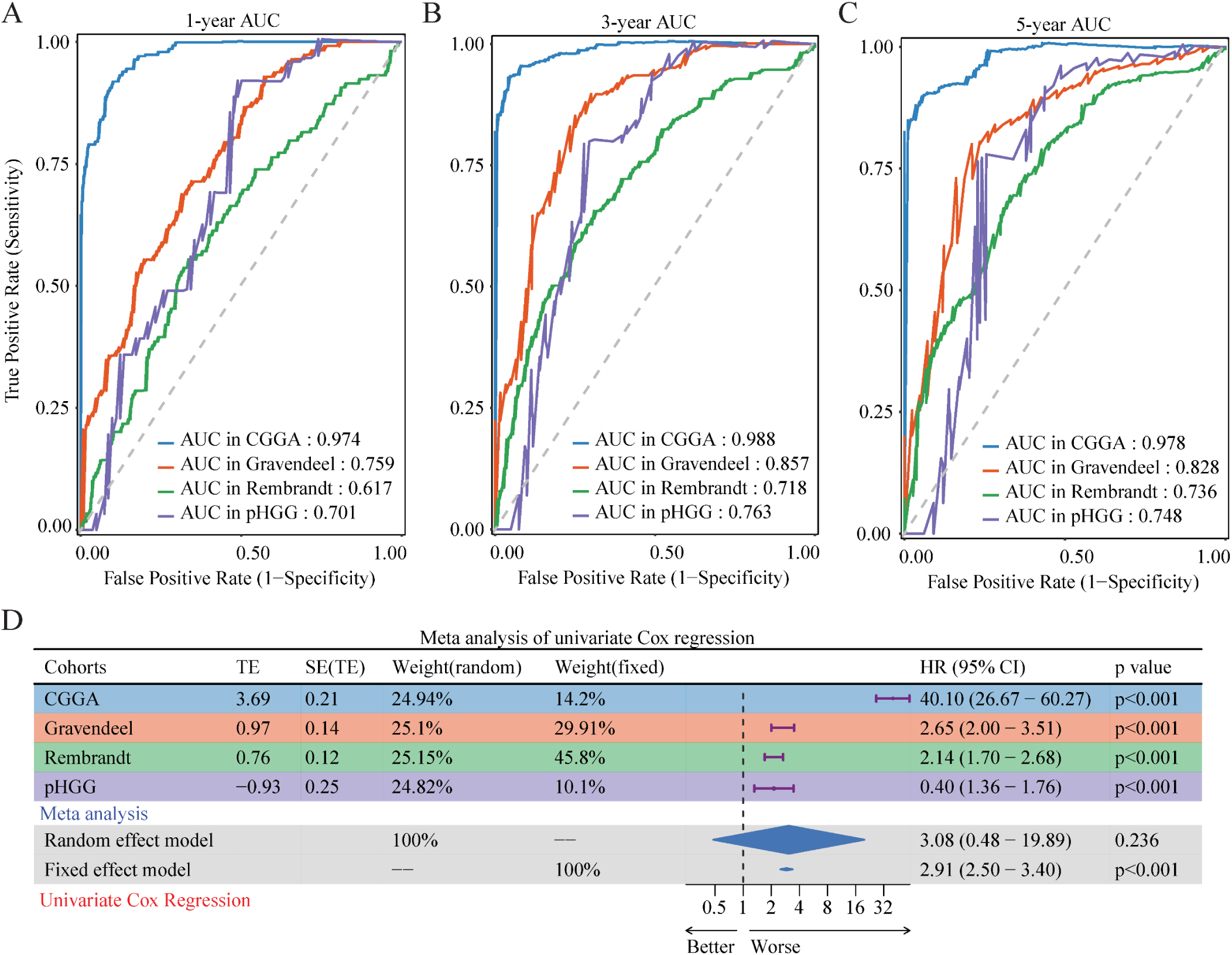
Validation of the final prognostic model. (A–C) Time-dependent ROC curves showing the predictive accuracy of the final StepCox[forward] + RSF model for overall survival at (A) 1 year, (B) 3 years, and (C) 5 years in the CGGA, Gravendeel, Rembrandt, and pHGG cohorts. The AUC for each cohort is indicated on the plots. (D) Forest plot summarizing a meta-analysis of the model’s prognostic value using univariate Cox regression. The analysis displays HRs and 95% CIs for each cohort, along with pooled HR estimates derived from both random-and fixed-effects models.

### 2.3. Integrated nomogram for prognostic prediction

To generate a clinically practical tool for individualized survival prediction, we first constructed a molecular signature through a systematic multi-step feature selection process. Using univariate Cox regression, iterative LASSO Cox analysis repeated 1,000 times, and forward stepwise selection, we identified five genes (*FAM86B1*, *IGFBP2*, *CA14*, *SCARA3*, and *TSPAN13*) with the most stable and significant prognostic value. The time-dependent ROC curve analysis revealed that the predictive performance of the model peaked when these five genes were included, achieving a 5-year AUC of 0.97 (Fig. 8A). Based on this model, a GSRScore was calculated for each patient. The prognostic accuracy of the GSRScore was further validated by ROC analysis, yielding an AUC of 0.973 (Fig. 8B). Patients were then stratified into high-and low-risk groups according to the median GSRScore, and Kaplan–Meier survival analysis confirmed a highly significant difference in overall survival between the two groups (log-rank p < 0.001; Fig. 8C), with high-risk patients exhibiting markedly poorer outcomes.

**Fig. 8.**
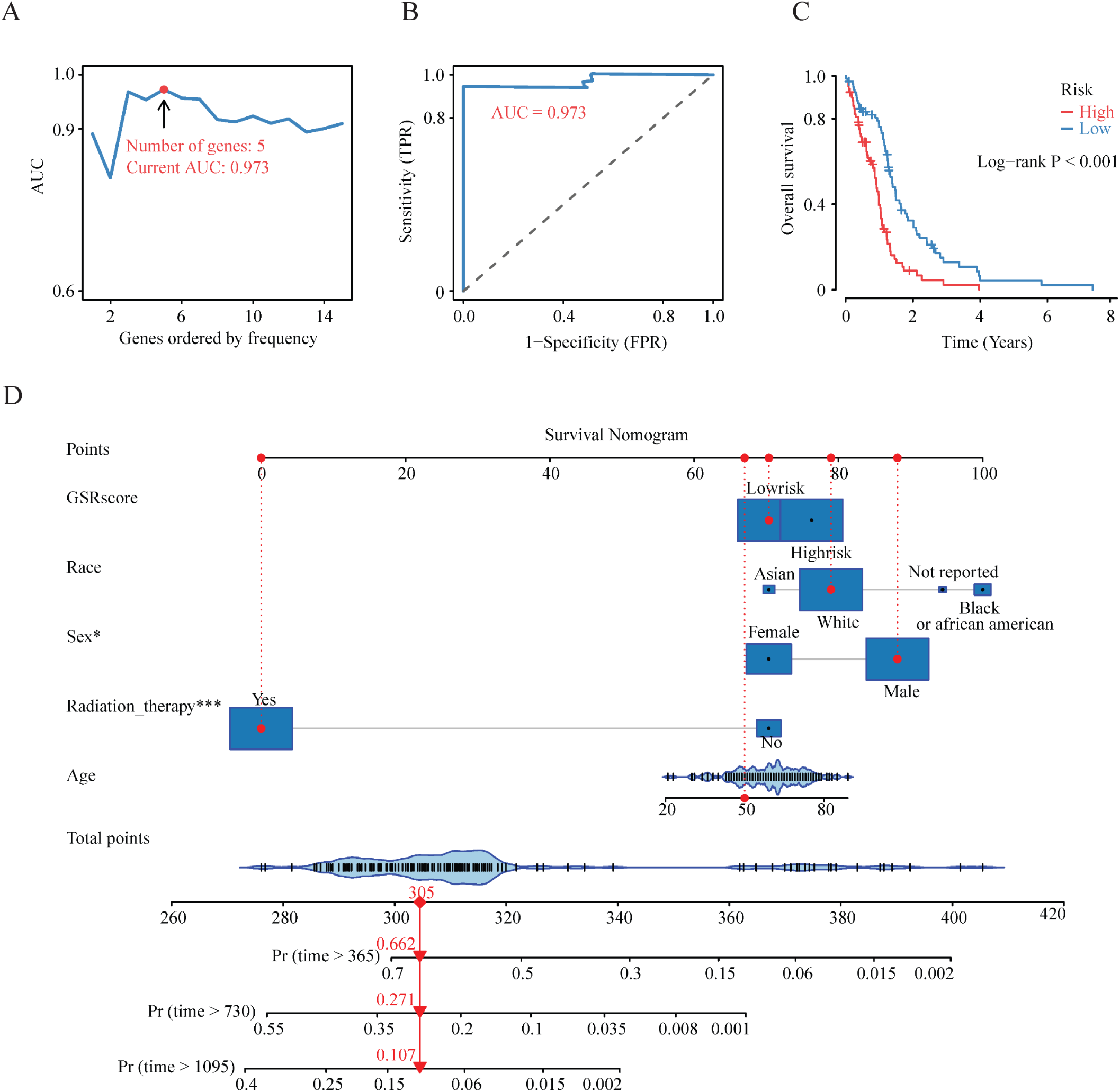
Development of a prognostic nomogram for clinical application. (A) Determination of the optimal number of genes for the prognostic risk signature based on the AUC. The model containing 8 genes achieved the highest predictive accuracy with an AUC of 0.99. (B) ROC curve analysis further validated the robust predictive performance of the 8-gene signature, yielding an AUC of 0.993. (C) Kaplan–Meier survival analysis demonstrating that the 8-gene signature provides significant prognostic stratification of HGG patients (Log-rank p < 0.001). (D) Construction of a prognostic nomogram integrating the gene signature risk score (GSRSscore) with independent clinical risk factors, including patient age, sex, race, and radiation therapy status. The nomogram was designed to predict 1-, 2-, and 3-year overall survival probabilities.

Building on this refined 5-gene signature, we next developed an integrated prognostic nomogram that combined the GSRScore with established clinical variables, including patient age, sex, race, and radiation therapy status (Fig. 8D). Each predictor was assigned a weighted point value proportional to its contribution to prognosis, and the total point score for an individual patient corresponded to predicted 1-, 2-, and 3-year overall survival probabilities. Collectively, this integrated nomogram provides a quantitative and user-friendly tool that improves prognostic accuracy and supports personalized risk assessment in patients with high-grade glioma.

### 2.4. FAM86B1-associated paralog axis supports glioma stemness

Among the five genes comprising the final prognostic signature incorporated into the nomogram, *FAM86B1* emerged as a particularly interesting candidate because of its limited prior characterization in glioma biology. Notably, FAM86B1 and FAM86B2 are closely related paralogs within the FAM86 family (13), a group of predicted methyltransferase-like proteins that share substantial sequence homology. Compared with the more full-length FAM86 family members, *FAM86B1* encodes a truncated FAM86 domain due to loss of a single exon, whereas *FAM86B2* more closely resembles the canonical family structure (Fig. 9A). Consistent with this relatedness, analysis of the public GBM dataset GSE54791 and our in-house DIPG13 RNA-seq dataset showed that FAM86B2, like FAM86B1, tended to be upregulated in GSCs relative to DGCs in most models, with the exception of the GBM MGG4 pair (Fig. 9B). In addition, higher FAM86B2 expression, similar to higher FAM86B1 expression, was associated with poorer GBM prognosis (Fig. 9C). Because the high sequence similarity between FAM86B1 and FAM86B2 complicates their selective detection and perturbation, we next examined the FAM86B1/2 axis across multiple patient-derived GSC models from both GBM and DIPG. Gene expression analysis demonstrated that FAM86B1/2 expression was elevated in GSCs relative to DGCs across GBM GSC lines as well as DIPG GSC models (Fig. 9D), supporting an association between this paralogous axis and the stem-like glioma state. To further assess clinical relevance, we examined FAM86B1/2 protein expression in human tissues by immunohistochemistry and found that GBM tumor specimens displayed higher expression than non-neoplastic tissues (Fig. 9E).

**Fig. 9.**
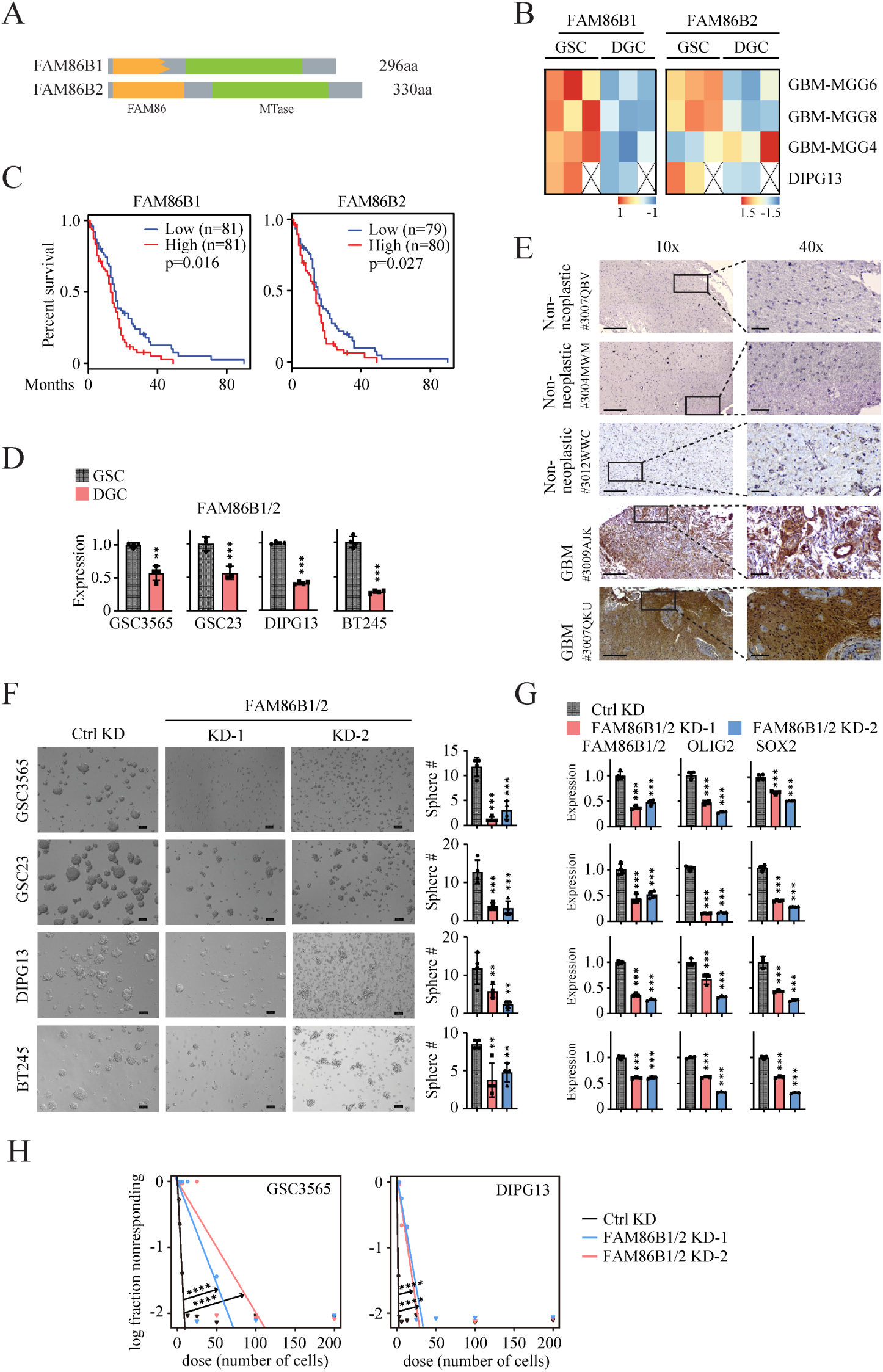
FAM86B1/2 axis supports glioma stemness. (A) Schematic comparison of human FAM86B1 and FAM86B2 protein domain architecture. FAM86B1 (296 amino acids (aa)) is shorter than FAM86B2 (330 aa) and encodes a truncated FAM86 domain. (B) Heatmaps showing FAM86B1 and FAM86B2 expression in GSCs and DGCs from the public GBM dataset GSE54791 and the in-house DIPG13 RNA-seq dataset. (C) Kaplan-Meier survival curves showing overall survival of patients with high versus low FAM86B1 or FAM86B2 expression in the TCGA GBM cohort generated using GEPIA2. Statistical significance was determined by log-rank test. (D) qPCR analysis of FAM86B1/2 expression in matched GSC and DGC models from GBM and DIPG. (E) Immunohistochemical analysis of FAM86B1/2 expression in human GBM and non-neoplastic brain tissues. Unpaired, two-tailed t test. Scale bars, 100 μm. (F) Representative images (left) and quantification (right) of tumorsphere formation in GSCs 72 hours after control or FAM86B1/2 knockdown. One-way ANOVA and Dunnett’s multiple-comparison test. Scale bars, 100 μm. (G) qPCR analysis of FAM86B1/2 as well as the stemness markers OLIG2 and SOX2 in control and FAM86B1/2 knockdown GSCs shown in panel F. One-way ANOVA and Dunnett’s multiple-comparison test. (H) Extreme limiting dilution analysis (ELDA) of GSC self-renewal following FAM86B1/2 knockdown. Data points represent the log fraction of wells without spheres plotted against the number of cells plated per well after 10 days. Statistical significance was determined by pairwise comparison. Data represent mean ± SD. **p < 0.01; ***p < 0.001; ****p < 0.0001.

We then performed functional knockdown studies using two independent MISSION shRNAs (MilliporeSigma) originally designed against FAM86B1. However, BLAST analysis revealed that both shRNAs also target FAM86B2, indicating likely co-suppression of both transcripts. In GBM and DIPG GSC models, shRNA-mediated suppression of the FAM86B1/2 axis reduced neurosphere formation capacity (Fig. 9F), decreased expression of the stemness regulators OLIG2 and SOX2 (Fig. 9G), and reduced self-renewal potential in limiting dilution assays (Fig. 9H), together indicating that the FAM86B1/2 paralogous axis contributes to maintenance of core GSC stemness programs. Collectively, these results provide experimental support for the biological relevance of a key gene family represented within our prognostic signature and strengthen the link between the GSC-associated transcriptomic program and functional stemness phenotypes in high-grade glioma.

## 3. Discussion

By contrasting patient-derived GSCs with their differentiated counterparts from both adult and pediatric HGGs, we defined a coherent set of 603 stemness-associated genes enriched in neurogenesis, RNA splicing, and oncogenic signaling pathways, underscoring their direct relevance to GSC biology. Network analysis highlighted SOX2, OLIG1, and OLIG2 as central regulators, consistent with their established roles in neural stem/progenitor cell identity and glioma plasticity (8, 14, 15). Accordingly, the risk score generated by our StepCox[forward] + RSF model likely quantifies the activity of this stemness program, with high-risk tumors reflecting a stronger GSC phenotype and poorer prognosis.

A key innovation of this study is the translation of this gene signature into a clinically usable nomogram. Although molecular risk scores are powerful, their direct adoption in practice is challenging. By integrating the GSRScore with clinical factors such as age, sex, race, and radiation therapy status, the nomogram provides an individualized prediction of 1-, 2-, and 3-year survival probabilities. This quantitative tool enables clinicians to move beyond population-level estimates and engage in more precise, personalized discussions with patients, supporting both clinical decision-making and risk stratification for future trials.

Beyond prognosis, our findings carry therapeutic implications. The GSC state is closely linked to resistance against standard-of-care treatments such as temozolomide and radiation, largely through enhanced DNA damage repair (DDR) (7, 16). It is plausible that patients classified as “high-risk” by our model represent those whose tumors are most refractory to conventional therapy. This opens opportunities for pairing the prognostic tool with therapeutic strategies. For instance, enrolling high-risk patients into clinical trials evaluating GSC-targeting agents or DDR inhibitors. In this way, the model could evolve into a predictive biomarker guiding precision therapy.

To further support the biological significance of the prognostic signature, we focused on *FAM86B1*, one of the five genes in the final model. Our experimental data suggest that FAM86B1 and its closely related paralog FAM86B2 are linked to the GSC state in both GBM and DIPG. Because the two shRNAs initially selected to target FAM86B1 were also found to target FAM86B2, the functional phenotypes observed here are best interpreted as resulting from suppression of the FAM86B1/2 axis rather than FAM86B1 alone. This interpretation is strengthened by the finding that FAM86B2, like FAM86B1, was upregulated in GSCs and associated with poor prognosis. The observed reduction in neurosphere formation together with decreased OLIG2 and SOX2 expression following FAM86B1/2 suppression supports a role for this paralogous axis in maintaining glioma stemness. More broadly, the fact that the other genes in the signature, including *IGFBP2* (17), *CA14* (18), *SCARA3* (19), and *TSPAN13* (20), have also been linked to poor prognosis in GBM further strengthens the biological and clinical relevance of the model.

Nevertheless, several limitations warrant consideration. First, our analyses were retrospective, and prospective, multi-center clinical trials will be essential to confirm real-world performance. Second, although the present study provides initial functional validation of the FAM86B1/FAM86B2 axis, the individual contributions of FAM86B1 and FAM86B2 remain to be fully resolved because of their high sequence similarity. Finally, tissue availability remains a barrier for HGG diagnostics. Future efforts may explore liquid biopsy approaches, such as circulating tumor DNA or extracellular vesicles, to detect the GSC signature non-invasively, which would greatly enhance clinical feasibility.

In conclusion, our integration of GSC transcriptomic profiling with machine learning, multi-cohort validation, and experimental interrogation of the FAM86B1/FAM86B2 axis produced a robust prognostic model for HGGs. By distilling complex biology into a clinically usable nomogram, we offer a practical tool for individualized survival prediction while reinforcing glioma stemness as a key driver of disease behavior and a promising target for therapeutic intervention.

## 4. Materials and methods

### 4.1. Cell culture

DIPG cells (DIPG13 and BT245) were maintained in tumor stem cell medium consisting of Neurobasal-A and DMEM/F-12 mixed 1:1 and supplemented with 10 mmol/L HEPES, sodium pyruvate, nonessential amino acids, GlutaMAX-I, and B27 (all from Thermo Fisher Scientific). Growth factors included human FGF (20 ng/mL), human EGF (20 ng/mL), and PDGF-AA and PDGF-BB (10 ng/mL each; FUJIFILM Biosciences), along with heparin (10 ng/mL; Stemcell Technologies). Under these conditions, DIPG cells grew as neurospheres, consistent with GSC properties. Patient-derived GBM GSCs, including GSC3565 and GSC23, were cultured according to established protocols (8). For differentiation, GSCs were maintained in DMEM/F-12 supplemented with 10% fetal bovine serum (FBS) for at least 14 days, resulting in DGCs that grew adherently. Lenti-X 293T cells (Takara Bio, cat# 632180) were cultured in DMEM (Gibco, cat# 11995065) supplemented with 10% FBS (Corning, cat# 35-015-CV).

### 4.2. Lentiviral-mediated gene knockdown

Lenti-X 293T cells were transfected using jetPRIME (Polyplus, cat# 101000001) with the packaging plasmids pMD2.G and psPAX2, together with either control shRNA or FAM86B1/2-targeting shRNAs (#1, TRCN0000131177; #2, TRCN0000130107) (Sigma-Aldrich). After 24 hours, the medium was replaced with GSC culture medium, and cells were incubated for an additional 48 hours before viral supernatants were collected and stored at −80°C. GBM and DIPG GSCs were infected with lentiviral supernatant for 18 hours, followed by medium replacement and incubation for an additional 48 hours before downstream analyses.

### 4.3. Tumorsphere assay and extreme limiting dilution assay (ELDA)

For the tumorsphere assay, GSCs expressing control or target shRNAs were dissociated into single cells and plated at 1 × 10^5 cells per well in 6-well plates under stem cell culture conditions. After 72 hours, tumorspheres were imaged using an Eclipse Ts2-FL microscope (Nikon) and quantified. To assess self-renewal frequency, the ELDA was performed. GSCs transduced with control or target shRNAs were dissociated into single cells and seeded into 96-well plates at serially diluted cell densities in triplicate under stem cell culture conditions. After 10 days, wells containing tumorspheres were scored, and stem cell frequency was estimated using the online ELDA tool (https://bioinf.wehi.edu.au/software/elda/).

### 4.4. Quantitative PCR (qPCR)

Total RNA was extracted using the RNeasy Plus Mini Kit (QIAGEN, cat# 74136) and reverse-transcribed using qScript cDNA SuperMix (Quantabio, cat# 95048). qPCR was performed in triplicate using PowerUp SYBR Green Master Mix (Applied Biosystems, cat# 25742) under the following cycling conditions: 95°C for 30 seconds, followed by 40 cycles of 95°C for 3 seconds and 60°C for 20 seconds. Primer sequences were as follows: FAM86B1/B2, forward 5′-ACTGTGAGGCATCCTGTGTG-3′ and reverse 5′-TGATGGCTGTGCTCTTGGAG-3′; OLIG2, forward 5′-TGGCTTCAAGTCATCCTCGTC-3′ and reverse 5′-ATGGCGATGTTGAGGTCGTG-3′; SOX2, forward 5′-GCCGAGTGGAAACTTTTGTCG-3′ and reverse 5′-GGCAGCGTGTACTTATCCTTCT-3′; and ACTIN, forward 5′-CATGTACGTTGCTATCCAGGC-3′ and reverse 5′-CTCCTTAATGTCACGCACGAT-3′.

### 4.5. Immunohistochemistry

Normal brain and GBM tissues (Supplementary Table 2) were obtained from the Enterprise Clinical Research Operations (ECRO) Biorepository at Indiana University Health Methodist Hospital with Institutional Review Board (IRB) approval for the collection and use of human biological materials. Tissues were processed as formalin-fixed, paraffin-embedded (FFPE) specimens, and immunohistochemical staining was performed as previously described (21). The primary antibody used for staining was anti-FAM86B1 (Thermo Fisher Scientific, cat# PA5-63164, 1:50). All slides were imaged using a Motic EasyScan Pro 6 scanner (Motic).

### 4.6. Data acquisition and preprocessing

To identify genes associated with the GSC phenotype, we analyzed both in-house and publicly available datasets. The in-house dataset was generated from DIPG13 cells, with total RNA extracted from GSC and DGC populations and sequenced by Novogene Corporation. In parallel, a publicly available GBM dataset (GSE54791; (22)) containing matched GSC and DGC pairs was included for comparative analysis.

For model construction and validation, one training cohort and three independent validation cohorts were assembled. The Chinese Glioma Genome Atlas (CGGA) RNA-seq dataset (http://www.cgga.org.cn/) served as the training cohort. Two well-established microarray datasets, Gravendeel (23) and Rembrandt (24), were used as validation cohorts. In addition, a pediatric HGG validation cohort was generated by integrating raw data from five high-quality studies: Hoffman (25), Paugh (26), Schwartzentruber (27), Sturm (28), and Witt (29). All data were uniformly curated and processed. Patients lacking complete survival information were excluded. RNA-seq data were normalized and log-transformed as log2(TPM+1) or log2(FPKM+1). Microarray data were processed from raw.CEL files using the Robust Multi-array Average (RMA) algorithm for background correction, normalization, and summarization. To ensure comparability across platforms, all expression values were standardized using z-scores prior to downstream analyses.

### 4.7. Identification of GSC–associated genes and pathway enrichment analysis

Differentially expressed genes (DEGs) between GSCs and DGCs were identified in both the in-house DIPG13 and GSE54791 datasets using the *limma* package (30) in R. Genes with |log2 fold change| > 1 and an adjusted p-value < 0.05 were considered significant. The intersection of DEGs from the two datasets yielded a set of 603 GSC–associated genes, which served as the candidate feature pool for model construction. Functional enrichment and network analyses were then performed to characterize these genes. A protein–protein interaction (PPI) network was constructed using STRING (https://string-db.org/) with a confidence score cutoff of 0.4 and visualized in Cytoscape (31). Hub modules were identified using the cytoHubba plugin (32). Gene Ontology (GO) (33) and Kyoto Encyclopedia of Genes and Genomes (KEGG) (34) enrichment analyses were carried out with *clusterProfiler* (35), with terms and pathways considered significant at adjusted p < 0.05.

### 4.8. Machine learning model construction and selection

The HGG GSC–associated genes served as input features for prognostic model development in the CGGA training cohort. A total of 135 modeling pipelines were constructed by pairing 10 feature selection strategies, including LASSO and stepwise Cox regression, with multiple survival learning algorithms such as Random Survival Forest (RSF), CoxBoost, and Supervised Principal Components (SuperPC). Model performance was assessed using the concordance index (C-index), a standard measure of survival prediction accuracy. The model with the highest mean C-index across all four cohorts was selected, and patient-specific risk scores were subsequently calculated based on this model.

### 4.9. Model validation and performance benchmarking

The final model was validated across all cohorts. Patients were stratified into high-and low-risk groups according to the median risk score derived from the training cohort. Survival differences between groups were assessed using Kaplan–Meier analysis with the log-rank test. Model discrimination was further evaluated using time-dependent Receiver Operating Characteristic (ROC) curves and the corresponding Area Under the Curve (AUC) values at 1-, 3-, and 5-year survival endpoints. To benchmark performance, the C-index and AUCs of our model were systematically compared against a broad panel of previously published glioma prognostic signatures across all four cohorts.

### 4.10. Meta-analysis

To evaluate the overall prognostic significance and generalizability of the model-derived risk score, a meta-analysis was performed across the four cohorts. Within each cohort, the risk score as a continuous variable was assessed using univariate Cox proportional hazards regression. Hazard ratios (HRs) with 95% confidence intervals (CIs) were pooled using a random-effects model based on the DerSimonian and Laird method (36), yielding a summary HR. This approach provided a robust estimate of prognostic value while accounting for heterogeneity among cohorts.

### 4.11. Construction and validation of a prognostic nomogram

A prognostic nomogram was constructed to provide individualized survival prediction by integrating molecular and clinical factors. A multivariable Cox proportional hazards regression model (37) was fitted using the cph function from the *rms* and *survival* packages in R (version 4.4.1), with overall survival time and status as dependent variables and the gene signature risk score (GSRScore), age, sex, race, and radiation therapy status as independent covariates. Based on the regression coefficients, a visual nomogram was generated using the *regplot* package, where each predictor was assigned a point value proportional to its contribution to prognosis. The total point score for each patient corresponded to predicted 1-, 2-, and 3-year survival probabilities. Model performance was assessed by discrimination and calibration, where discrimination was quantified by the C-index (0.5 indicates no discrimination and 1.0 indicates perfect discrimination), and calibration was evaluated by comparing predicted and observed survival probabilities using calibration curves generated from 1,000 bootstrap resamples to reduce overfitting.

### 4.12. Statistical analysis

All statistical analyses were performed in R. Survival analyses were conducted using the *survival* (https://CRAN.R-project.org/package=survival) and *survminer* (https://CRAN.R-project.org/package=survminer) packages. Time-dependent ROC analyses were carried out with the *timeROC* package (38), and meta-analyses were implemented using the *meta* package (39). A two-sided p value < 0.05 was considered statistically significant.

### 4.13. Data availability

RNA-seq data for DIPG13 cells cultured under stem cell-maintaining and differentiation conditions will be deposited in the NCBI Gene Expression Omnibus (GEO). All other data supporting the findings of this study are available from the corresponding authors upon reasonable request.

## Author contributions

QX: Investigation, Visualization, Data curation, writing–original draft. BW: Investigation, Visualization, Data curation, writing–original draft. QM: Writing–review and editing. JS: Conceptualization, Methodology, Funding acquisition, Supervision, Project administration, Writing–original draft, Writing–review and editing.

## Data Availability

All data produced in the present study are available upon reasonable request to the authors

https://www.ncbi.nlm.nih.gov/geo/

## Acknowledgments

This study was supported by the ACS-IRG Grant Mechanism (22-147-37; J. Shen), the Schwarz Family and Friends Cancer Research Fund (J. Shen), the Indiana University Seed Grant (J. Shen), the IUSCCC Early Career Investigator Pilot Fund (J. Shen), and the IUSM LAMP PLUS Scholars Project Fund (J. Shen). The authors thank Dr. Jeremy Rich at the University of North Carolina for providing the GBM GSC lines. The authors also thank Dr. Stefanie Galban and Dr. Sriram Venneti at the University of Michigan, Dr. Q. Richard Lu at the University of Cincinnati, and Dr. Michelle Monje at Stanford Medicine for providing the DIPG cell lines.

